# Cost Minimisation and Threshold Analysis of Anatomical Endoscopic Enucleation of the Prostate

**DOI:** 10.64898/2026.08.15.26360519

**Authors:** Julene Ong Hui Wun, Rachel Lau Shu En, Kit Mun Chow, Daanesh Huned, Roxanne Teo Yong Ai, Han Jie Lee, Ee Jean Lim, Edwin Jonathan Aslim, Yong Wei Lim, Kelven Chen, Yi Quan Tan, Joon Jae Park, Joshua Tung Yi Min

## Abstract

**Introduction:** Anatomical endoscopic enucleation of the prostate (AEEP) techniques, including bipolar enucleation (B-TUEP), holmium laser enucleation (HoLEP), thulium laser enucleation (ThuLEP), and thulium fibre laser enucleation (ThuFLEP), demonstrate comparable clinical outcomes for benign prostatic hyperplasia. As clinical equivalence is increasingly established, cost becomes a key determinant of modality selection. We performed a cost minimisation analysis comparing index procedural costs across AEEP modalities from an institutional perspective.

**Methods:** A cost minimisation model was developed from the institutional perspective, incorporating amortised capital costs, maintenance, and consumables. In addition to the base-case scenario of 180 cases per year, we modelled two additional case volume scenarios: low (50 cases/year) and high (500 cases/year) volume. Thu:YAG laser fibres were modelled on two scenarios: disposable single-use, and reusable fibres (up to 10 cases per fibre). Breakeven analysis determined the threshold volume at which each laser modality achieves cost parity with B-TUEP, and one-way sensitivity analysis was performed on key cost parameters. Analysis was limited to index procedural costs calculated in Singapore dollars.

**Results:** At the base case of 180 cases per year, B-TUEP had the lowest index procedure cost (SGD 1,018), followed by ThuFLEP (SGD 1,584), ThuLEP (1,599), and HoLEP (SGD 1,655). Breakeven analysis demonstrated that HoLEP, ThuLEP, and ThuFLEP can never achieve cost parity with B-TUEP when laser fibres are single-use, as laser modalities carry higher costs on both capital and per-case dimensions. ThuLEP with reusable fibres (10 uses per fibre) was the only modality to cross below B-TUEP, at a breakeven volume of 198 cases per year. At 500 cases per year with reusable fibres, ThuLEP achieved the lowest cost (SGD 847), representing a 15.4% saving over B-TUEP. Sensitivity analysis identified annual case volume and B-TUEP loop cost as the most influential parameters.

**Conclusion:** Index procedural costs in AEEP are strongly influenced by case volume and consumable strategy. While B-TUEP remains cost-efficient at low volume, high-volume practice combined with reusable Thu:YAG fibre technology enables cost parity and potential cost advantage for laser enucleation. These findings highlight the importance of economies of scale and device utilisation in technology adoption.

## Introduction

Anatomical endoscopic enucleation of the prostate (AEEP) has gained increasing acceptance as a size-independent alternative to transurethral resection (TURP) and open simple prostatectomy for the surgical management of benign prostatic hyperplasia (BPH), with particular advantages for large-volume prostates (1). Multiple energy sources are now available for enucleation, including bipolar electrosurgery (B-TUEP), holmium:YAG lasers (HoLEP), pulsed thulium:YAG lasers (ThuLEP), and thulium fibre lasers (ThuFLEP). While the evidence base for newer energy modalities such as ThuFLEP is still maturing, available trials and meta-analyses suggest broadly comparable efficacy and safety profiles across AEEP techniques (2–5).

Given clinical equivalence, the selection of an AEEP platform increasingly depends on factors such as equipment cost, operative logistics, and institutional infrastructure. However, the cost structures of these modalities differ substantially. B-TUEP utilises a conventional bipolar generator, which is inexpensive and widely available. Laser-based modalities require dedicated laser platforms, which can cost between SGD 188,000 and SGD 320,000, but may offer advantages in consumable reuse, as some laser fibres can be reprocessed for multiple uses (6). Previous cost analyses of endoscopic prostate surgery have largely focused on comparisons between enucleation and resection (TURP), and direct cost comparisons across AEEP modalities remain sparse. Notably, no study has examined the interaction between case volume and fibre reuse strategy as determinants of cost competitiveness among AEEP techniques.

We conducted a cost minimisation analysis comparing the index procedural costs of four AEEP modalities (B-TUEP, HoLEP, ThuLEP, and ThuFLEP) across varying annual case volumes. A breakeven analysis was performed to determine the threshold volume at which each laser modality achieves cost parity with B-TUEP, and one-way sensitivity analysis was used to identify the most influential cost parameters.

## Methods

### Study Design & Perspective

This was a micro-costing study conducted from the institutional (hospital) perspective. We developed a deterministic cost model comparing the per-case index procedure cost of four AEEP modalities at high-volume tertiary academic medical centres in Singapore. The model uses Singapore dollar (SGD) costs. As the analysis involved no patient-level data, Institutional Review Board approval was not required.

### Cost Components

The total index procedure cost was defined as the sum of amortised capital equipment cost, per-case consumables, energy delivery device cost, anaesthesia cost, and surgeon fee. Capital equipment costs were obtained from institutional procurement records and amortised over an assumed useful life of 8 years. Annual maintenance was estimated at 6% of the capital purchase price based on institutional service contract data. The annualised capital cost per case was calculated as:

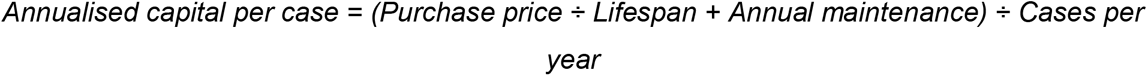

Consumables included morcellator blades, irrigation tubing, and catheters. Energy delivery costs comprised bipolar loops (B-TUEP) or laser fibres (HoLEP, ThuLEP, ThuFLEP). In the base case, all energy delivery devices were modelled as single-use. A separate scenario modelled ThuLEP fibre reuse (up to 10 procedures per fibre), reflecting the reprocessing capability of pulsed-thulium laser fibres.

Costs common to all modalities, including operating theatre time, surgical and anesthetic fees, nursing staff, monitoring equipment, and perioperative consumables (saline, chlorhexidine, gauzes) were excluded, as they do not differ between modalities and cancel in a cost minimisation framework. Inpatient costs (ward charges, transfusion costs) were also assumed equivalent across modalities based on available evidence (5,7) and institutional practices, where AEEP patients follow the same postoperative protocols, and were therefore excluded from the primary analysis.

### Volume Scenarios

Three annual case volume scenarios were modelled to reflect varying institutional throughput: low volume (50 cases per year), base case (180 cases per year, reflecting our institutional caseload for prostates ≥30 g undergoing surgery), and high volume (500 cases per year). The same capital equipment, maintenance, and consumable costs were applied across all scenarios; only the denominator for capital cost amortisation changed.

### Breakeven Analysis

For each laser modality, the breakeven volume - the annual case volume at which the per-case cost equals that of B-TUEP - was derived algebraically. The total cost per case can be decomposed into a volume-dependent component (annualised fixed costs divided by case volume) and a volume-independent component (per-case variable costs):

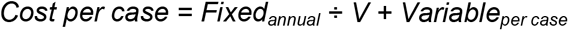

Setting the laser cost equal to the B-TUEP cost and solving for volume (V) yields:

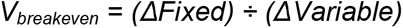

Where Δ*Fixed* = Laser annual fixed cost - B-TUEP annual fixed cost, and Δ*Variable* = B-TUEP variable cost - Laser variable cost. A positive breakeven volume exists only when B-TUEP has higher per-case variable costs than the comparator laser modality (i.e., the denominator is positive). If both fixed and variable costs are higher for the laser, no breakeven exists at any volume.

### Sensitivity Analysis

A one-way deterministic sensitivity analysis was performed on eleven parameters: annual case volume (50-500), capital equipment costs (±20-30% of base case for each modality), equipment lifespan (5-12 years), maintenance rate (3-10% of capital), laser fibre costs (±30%), ThuLEP fibre reuse count (5-10 uses), and B-TUEP loop cost (SGD 200-450). Each parameter was varied independently while all others were held at base-case values. Results are reported as the change in cost difference versus B-TUEP at the base-case volume of 180 cases per year.

## Results

### Base-case Analysis

**Table 1** presents the cost breakdown by modality at the base-case volume of 180 cases per year. B-TUEP had the lowest total index procedure cost (SGD 1,018), primarily driven by its substantially lower capital equipment cost (SGD 25,862 vs SGD 188,000-320,000 for laser platforms). Among the laser modalities, ThuFLEP was the least expensive (SGD 1,584), followed by ThuLEP (SGD 1,599) and HoLEP (SGD 1,655). The cost difference between B-TUEP and the laser modalities ranged from SGD 566 (ThuFLEP) to SGD 637 (HoLEP), corresponding to a 55.6-62.5% premium for laser enucleation.

**Table 1.** Index procedure cost breakdown by AEEP modality (base case: 180 cases/year). Costs in SGD.

| Cost Component | B-TUEP | HoLEP | ThuLEP | ThuFLEP |
| --- | --- | --- | --- | --- |
| Capital equipment | 25,862 | 320,000 | 300,000 | 188,000 |
| Annualised capital/case | 26.58 | 328.89 | 308.33 | 193.22 |
| Consumables/case | 691.38 | 640.80 | 640.80 | 640.80 |
| Energy delivery/case | 300.00 | 685.00 | 650.00 | 750.00 |
| **Total index procedure cost** | **1,018** | **1,655** | **1,599** | **1,584** |
| *Difference vs B-TUEP* | *Ref* | +637 | +581 | +566 |

### Volume Scenario Analysis

Table 2 summarises the total index procedure cost across the three volume scenarios. At low volume (50 cases per year), the cost advantage of B-TUEP was most pronounced; B-TUEP cost SGD 1,087 compared with SGD 2,510 for HoLEP, SGD 2,401 for ThuLEP, and SGD 2,086 for ThuFLEP, representing a 91.9-130.9% cost advantage. At high volume (500 cases per year), per-case costs decreased across all modalities, but B-TUEP remained the least expensive (SGD 1,001) when laser fibres were single-use. The cost gap narrowed to 40-45.9%, as the volume-dependent capital component became proportionally smaller.

**Table 2.** Index procedure cost by volume scenario (SGD). ThuLEP (R) = reusable fibre, 10 uses.

| Volume | B-TUEP | HoLEP | ThuLEP | ThuLEP (R) | ThuFLEP |
| --- | --- | --- | --- | --- | --- |
| 50/yr | 1,087 | 2,510 | 2,401 | 1,846 | 2,086 |
| 180/yr | 1,018 | 1,655 | 1,599 | 1,044 | 1,584 |
| 500/yr | 1,001 | 1,444 | 1,402 | 847 | 1,460 |

The volume sensitivity was markedly asymmetric across modalities. Increasing volume from 50 to 500 cases per year reduced HoLEP costs by 42.4% (SGD 2,510 to SGD 1,444), ThuLEP by 41.6% (SGD 2,401 to SGD 1,402), and ThuFLEP by 30% (SGD 2,086 to SGD 1,460), but reduced B-TUEP costs by only 7.9% (SGD 1,087 to SGD 1,001). This asymmetry reflects B-TUEP’s low capital cost, which makes its per-case cost relatively volume-insensitive.

### Breakeven Analysis

The breakeven analysis showed that when laser fibres are used as single-use disposables, no laser modality can achieve cost parity with B-TUEP at any volume. This is because all three laser modalities carry higher costs than B-TUEP on both the volume-dependent dimension (higher capital expenditure) and the volume-independent dimension (more expensive energy delivery disposable per case). Increasing volume reduces the capital cost gap, but cannot eliminate the per-case variable cost disadvantage.

ThuLEP with reusable fibres was the sole exception. Fibre reuse (10 uses per fibre) reduces the per-case energy delivery cost from SGD 650 to SGD 95, making ThuLEP’s variable cost (SGD 736) lower than B-TUEP’s (SGD 991). This creates a positive denominator in the breakeven equation, yielding a breakeven volume of 198 cases per year. Above this threshold, ThuLEP with reusable fibres becomes progressively cheaper than B-TUEP (**Figure 1**).

**Figure 1.**
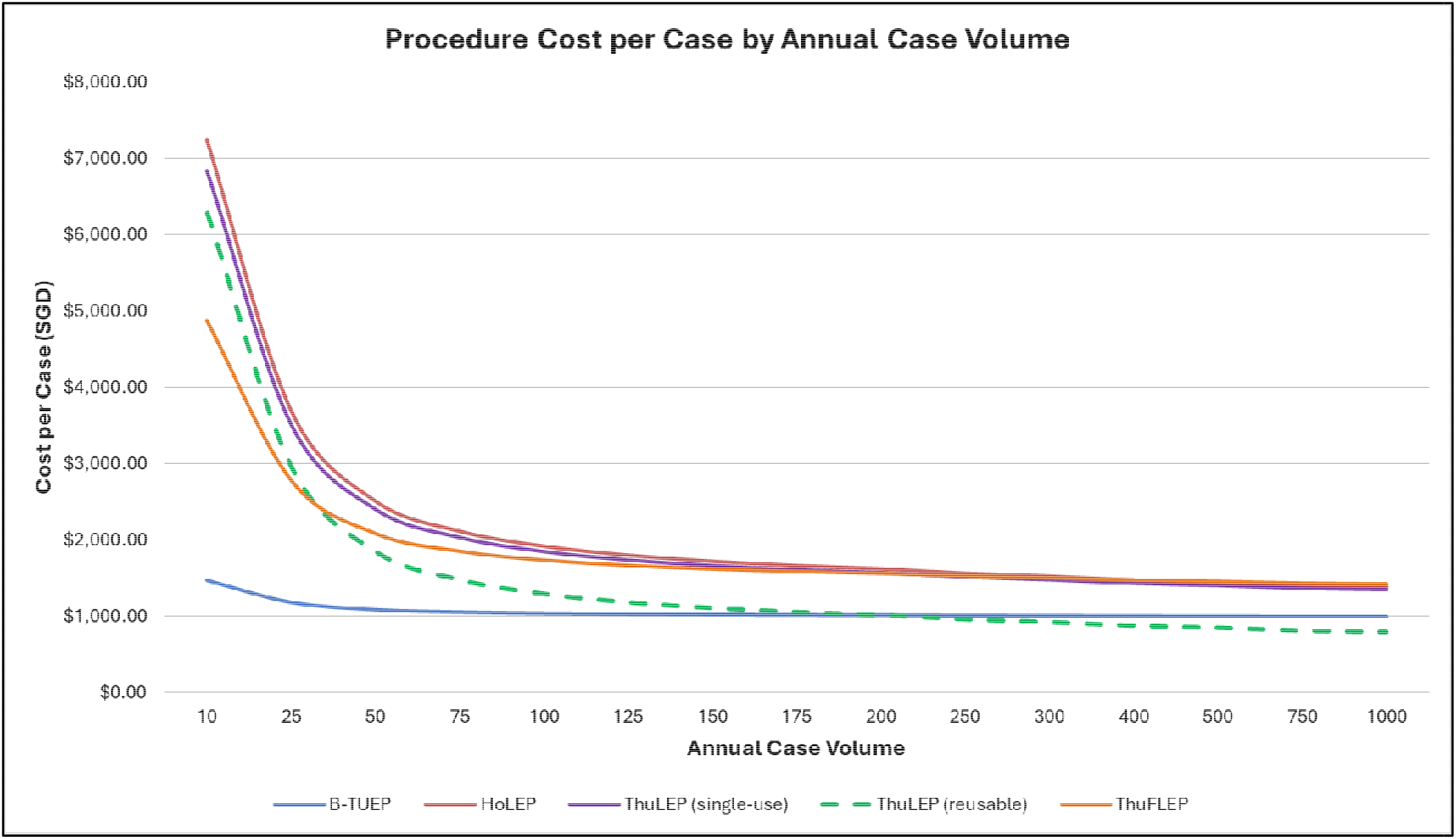
Cost per case versus annual case volume for all AEEP modalities. The ThuLEP (reusable) curve crosses below B-TUEP at approximately 198 cases per year.

### Sensitivity Analysis

The one-way sensitivity analysis for ThuLEP (reusable) versus B-TUEP at the base-case volume of 180 cases per year is presented in **Figure 2**. The parameters with the greatest influence on the cost difference were annual case volume, B-TUEP loop cost, and ThuLEP capital equipment cost. Equipment lifespan, maintenance rate, and fibre reuse count had moderate effects.

**Figure 2.**
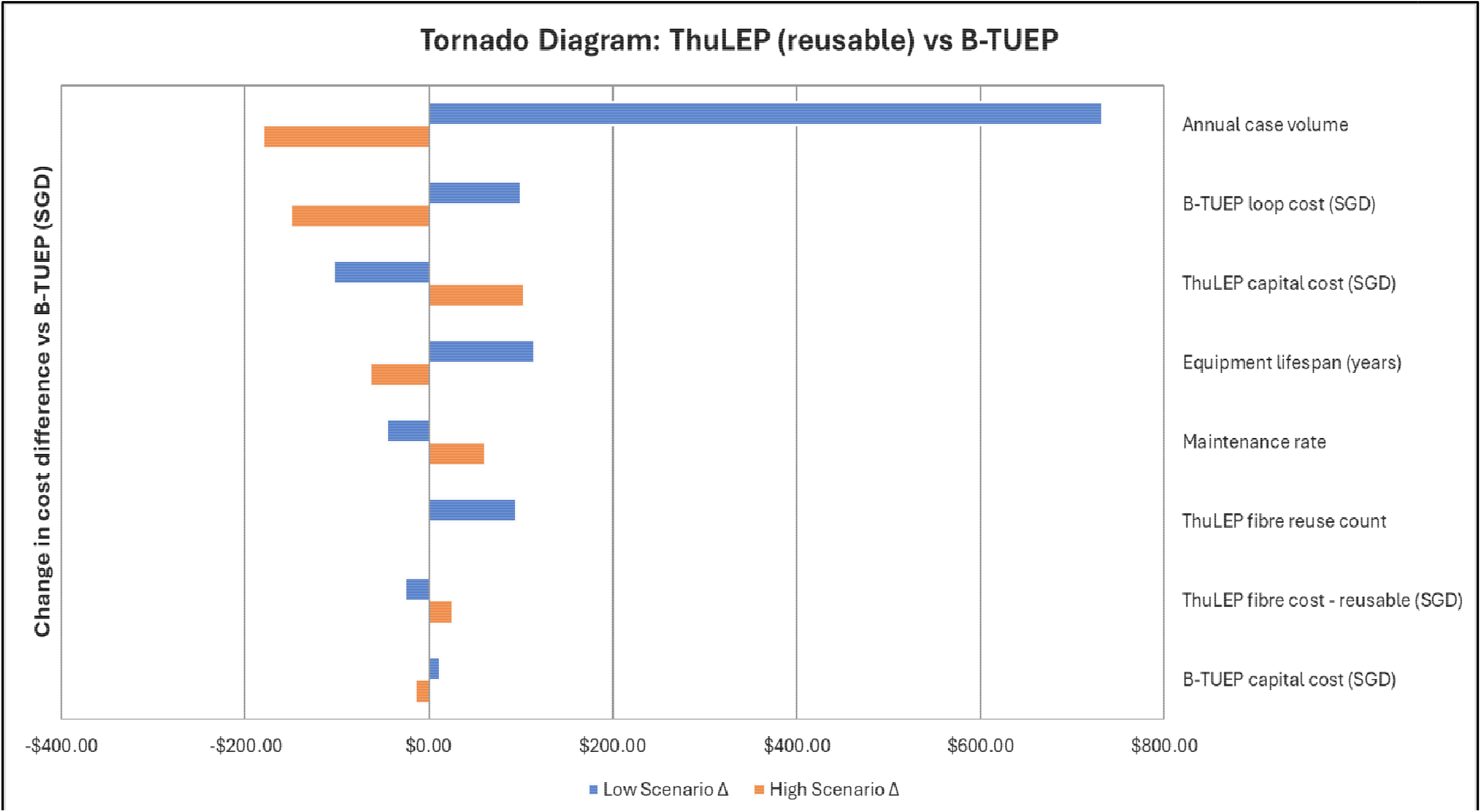
Tornado diagram showing one-way sensitivity analysis for ThuLEP (reusable) versus B-TUEP cost difference.

## Discussion

Several prior studies have examined costs associated with prostate enucleation, but to date no study has compared costs across three or more enucleation energy sources in a formal economic evaluation. Wymer et al. developed a Markov cost-effectiveness model comparing HoLEP with B-TURP, WVTT, and PUL, finding HoLEP most cost-effective, but did not compare across enucleation modalities (8). Higazy et al. provided the only direct cost comparison between two AEEP techniques (HoLEP vs B-TUEP), with a cost advantage of HoLEP over B-TUEP driven by earlier catheter removal and shorter hospital stay in the HoLEP group (9). This difference was attributed to the superior intraoperative hemostasis of HoLEP, but may have also been influenced by institutional practice - Bhandarkar et al. found no differences in bladder irrigation time, catheterization time, or hospital stay between HoLEP and B-TUEP in a prospective RCT (10). Likewise, a meta-analysis of 31 studies by Pallauf et al. found no differences in post-operative complications or catheterization time between enucleation modalities (11).

This study provides the first direct cost comparison across four AEEP modalities incorporating volume-dependent breakeven analysis and sensitivity analysis. Our principal finding is that B-TUEP is the most cost-efficient AEEP modality under most realistic conditions. When laser fibres are used as single-use devices, no laser modality can achieve cost parity with B-TUEP regardless of institutional volume. The only scenario in which a laser modality becomes cost-competitive is with reusable fibres at volumes exceeding ∼198 cases per year.

Our findings have direct implications for institutional procurement decisions. The 12-fold difference in capital equipment cost between B-TUEP (SGD 25,862) and HoLEP (SGD 320,000) represents a significant acquisition barrier, particularly for centres in resource-constrained settings or those initiating an AEEP programme. Our analysis demonstrates that this capital cost disadvantage can be overcome by ThuLEP when fibre reuse is practised - a strategy which may become available to HoLEP or ThuFLEP, if the associated laser fibres are multi-use.

We found that B-TUEP’s cost is relatively insensitive to volume (7.9% reduction from 50 to 500 cases per year), because its low capital cost means the volume-dependent amortisation component is small. In contrast, laser modalities show 30-42.4% cost reductions over the same volume range. This asymmetric volume-dependent behaviour of per-case costs is of practical importance, as B-TUEP is particularly advantaged in low-volume settings, where the capital cost burden per case is highest for laser platforms.

Our breakeven analysis also explains why cost comparisons that examine only a single volume scenario can be misleading. At high volume, laser modalities appear to approach parity with B-TUEP, but this convergence is asymptotic, as the per-case variable cost difference ensures that full parity is never reached without fibre reuse. We would encourage future cost analyses to present volume-dependent curves rather than single-point estimates.

### Limitations

This study has several limitations. First, the analysis is limited to index procedure costs and does not incorporate downstream costs such as complication management, follow-up, or re-intervention. However, as current evidence points towards clinical outcome equivalence across AEEP modalities, these costs are unlikely to differ meaningfully between arms. Should future studies demonstrate clinically significant differences in outcomes across modalities, a cost-effectiveness analysis incorporating long-term costs and quality-adjusted life years would be warranted, and the conclusions of this cost-minimisation framework would need to be revisited.

Second, equipment costs reflect institutional procurement at a single centre and may vary by region, negotiated contracts, and distributor-specific pricing. The sensitivity analysis partially addresses this uncertainty. Third, we did not account for opportunity costs of equipment downtime, training, or learning curve effects. Fourth, equipment sharing across procedures (e.g., a holmium laser used for both enucleation and lithotripsy) would reduce the effective capital cost allocated to AEEP cases; our model conservatively allocates the full capital cost to the AEEP programme.

## Conclusion

In a cost minimisation analysis of four AEEP modalities from an institutional perspective, B-TUEP was the most cost-efficient technique under the majority of realistic scenarios. Laser enucleation modalities carry a capital cost premium that cannot be offset by volume alone when fibres are single-use. ThuLEP with reusable fibres is currently the only modality that achieves cost parity with B-TUEP, at a threshold of approximately 198 cases per year. These findings provide an evidence-based framework for guiding procurement decisions and resource allocation in AEEP programme development.

## Data Availability

The datasets generated and analysed during the current study are not publicly available because they contain institution-specific procurement and pricing information that are commercially sensitive. De-identified data supporting the findings of this study may be available from the corresponding author on reasonable request, subject to institutional approval.

## Declarations

### Abbreviations

AEEP: Anatomical endoscopic enucleation of the prostate
BPH: Benign prostatic hyperplasia
B-TUEP: Bipolar transurethral enucleation of the prostate
HoLEP: Holmium laser enucleation of the prostate
ThuLEP: Thulium laser enucleation of the prostate
ThuFLEP: Thulium fibre laser enucleation of the prostate
SGD: Singapore dollar

### Ethics approval and consent to participate

As this study was a cost-minimisation analysis based on institutional procurement data and did not involve human participants, patient data, or biological specimens, ethics approval and informed consent were not required.

### Consent for publication

Not applicable.

### Authors’ contributions

Julene Ong Hui Wun contributed to the conceptualization of the study, data curation, investigation, formal analysis, manuscript drafting, and manuscript revision. Rachel Lau Shu En, Chow Kit Mun, Daanesh Huned, and Roxanne Teo Yong Ai contributed to data collection and manuscript drafting. Joshua Tung Yi Min contributed to study conceptualization, methodology, formal data analysis, and critical revision of the manuscript. Lee Han Jie, Lim Ee Jean, Edwin Jonathan Aslim, Lim Yong Wei, Kelven Chen, Tan Yi Quan, and Park Joon Jae provided supervision, expert guidance, and critical review of the manuscript. All authors read and approved the final manuscript.

## Acknowledgements

Not applicable

## Statements and Declarations

The authors have no conflicts of interest to declare.

No funding was received for this work.

As this study does not involve any new research with human participants, formal ethics approval was not required.

